# Factors Influencing High-Risk Fertility Practices Among Women of Reproductive Age in Zambia: A Multilevel Analysis of the 2024 Demographic and Health Survey

**DOI:** 10.64898/2026.01.23.26344686

**Authors:** Samuel Mutasha, Chanda Nkandu, Given Moonga

## Abstract

**Background:** High-risk fertility behaviors (HRFB) remain a public health concern in Zambia due to their association with maternal and newborn complications. Despite improvements in reproductive health services, updated national evidence on the magnitude and determinants of HRFB is limited. This study used data from the 2024 Zambia Demographic and Health Survey (ZDHS) to estimate the prevalence of HRFB and identify associated factors among women of reproductive age.

**Methods:** A cross-sectional analysis was conducted using weighted data from 13,951 women aged 15–49 years. HRFB was defined as experiencing at least one of the following: first birth before 18 years, last birth after 34 years, short birth interval (<24 months), or high parity (≥4 births). Explanatory variables included socio-demographic, economic, and reproductive health factors. Multilevel mixed-effects Poisson regression with robust standard errors was used to estimate adjusted prevalence ratios (aPR) and 95% confidence intervals (CI), accounting for clustering at the community level. Four models were fitted, and model fit was assessed using deviance, AIC, BIC, intra-class correlation (ICC), and median odds ratio (MOR).

**Results:** The prevalence of HRFB was 48.4%. Older maternal age increased risk (25–34 years: aPR 1.28; 95% CI 1.20–1.37; 35–49 years: aPR 1.89; 95% CI 1.78–1.99), while higher maternal and partner education were protective (secondary: aPR 0.78; tertiary: aPR 0.53). Employment and contraceptive use were associated with higher HRFB, whereas prior pregnancy termination and lower household wealth were protective. Rural residence was not significant. Minimal community-level variation was observed (ICC ≈ 0, MOR = 1), indicating that most variation occurred at the individual level.

**Conclusion:** HRFB remain highly prevalent in Zambia, with older maternal age, lower education, employment, and contraceptive use after risk onset as key risk factors. Interventions should prioritize improving education, promoting early family planning uptake, and supporting older women continuing childbearing to reduce HRFB and improve maternal and newborn outcomes.

## Introduction

High-risk fertility behaviors (HRFB) remain a critical and persistent public health challenge in sub-Saharan Africa, a region that continues to bear a disproportionate burden of global maternal and neonatal morbidity and mortality. HRFB commonly defined as early childbearing (<18 years), advanced maternal age (>34 years), short birth intervals (<24 months), and high parity (≥4 births) are consistently associated with adverse maternal and perinatal outcomes, including obstetric complications, preterm delivery, low birth weight, and elevated neonatal mortality [1], [2], [3]. These fertility patterns reflect underlying structural and social vulnerabilities, including limited access to reproductive health services, low educational attainment, and entrenched gender inequalities. As such, the persistence of HRFB poses a substantial barrier to progress toward the Sustainable Development Goals (SDGs), particularly those targeting reductions in maternal mortality and preventable neonatal deaths.

Despite substantial global declines in fertility over recent decades, sub-Saharan Africa continues to experience the highest burden of HRFB and their associated adverse maternal and child health outcomes. The region’s total fertility rate (TFR) was estimated at 4.7 births per woman during 2015–2020 more than twice the global average and markedly higher than that of any other world region[4]. Persistently high fertility in sub-Saharan Africa is closely intertwined with elevated maternal mortality, with the region recording an estimated maternal mortality ratio (MMR) of 546 deaths per 100,000 live births, representing both the highest global burden and the slowest rate of decline worldwide [4].

Within the region, the prevalence of HRFB remains strikingly high and heterogeneous across countries and population subgroups. A pooled analysis of nine East African countries reported that 57.6% of women of reproductive age experienced at least one HRFB, with pronounced disparities between urban and rural settings [3]. Country-specific estimates indicate even higher burdens; in Tanzania, for instance, 71.6% of women were reported to have at least one HRFB [5]. The consequences of these fertility patterns extend beyond maternal health, contributing substantially to poor child growth and survival. Across 32 sub-Saharan African countries, the prevalence of stunting among children under five a key adverse outcome associated with HRFB was estimated at 31.3%, alongside wasting at 8.1% and underweight at 17.0%[6].

In Zambia, fertility levels remain among the highest globally, with a total fertility rate (TFR) of 5.5 births per woman as of 2015 [7]. Marked rural–urban disparities persist, with women in rural areas averaging 6.6 births compared with 3.7 among urban women [7]. These fertility patterns intersect with suboptimal child health outcomes: national estimates indicate that among children under five years of age, 17.7% are stunted, 9.3% are underweight, and 5.6% are wasted outcomes that have been consistently linked to HRFB [6]. Although modern contraceptive use among married women has increased to 45%, coverage remains substantially lower in rural areas (39%) than in urban areas (53%), and unmet need for family planning continues to be disproportionately high in rural settings (24% versus 17%) [7].

Despite notable progress in expanding reproductive, maternal, and child health services, HRFB remain widespread in Zambia and continue to contribute to persistently elevated maternal and neonatal mortality [8], [9], [10]. Successive rounds of the Zambia Demographic and Health Survey (ZDHS) document high levels of early marriage, adolescent childbearing, short birth intervals, and high parity risk factors that act synergistically to heighten the likelihood of adverse reproductive and perinatal outcomes[1], [8], [10]. These behaviors are shaped by a complex constellation of socioeconomic and demographic factors, including educational attainment, household wealth, rural residence, marital status, cultural norms, and access to family planning services [1], [7], [11]. Importantly, the distribution and determinants of HRFB are dynamic, varying across population subgroups and over time, underscoring the need for updated, context-specific evidence.

The release of the 2024 Zambia Demographic and Health Survey provides a timely opportunity to generate contemporary, nationally representative evidence on the prevalence and determinants of HRFB. Such evidence is essential for informing targeted interventions aimed at populations at highest risk including adolescents, rural women, and women with limited formal education—and for accelerating progress toward reducing preventable maternal and neonatal deaths [12], [13], [14]. However, despite the recognized public health importance of HRFB, recent empirical analyses using up-to-date national data remain limited, particularly those that examine the multilevel interplay of individual, household, and contextual factors influencing women’s fertility behaviors in Zambia.

This study therefore examined the prevalence and determinants of HRFB among women of reproductive age in Zambia using data from the 2024 DHS, applying multilevel analysis to account for clustering at household and community levels. By generating evidence on both individual and contextual drivers of HRFB, this study seeks to inform targeted reproductive health interventions, strengthen family planning strategies, and support national efforts to reduce preventable maternal and neonatal mortality [15], [16].

## Method

### Study design, data source, and setting

This study used data from the 2024 Zambia Demographic and Health Survey (ZDHS), a nationally representative cross-sectional survey employing a stratified, multistage sampling design. The data were obtained from the DHS Program website on November 3, 2025. In the first stage, clusters or Enumeration Areas (EAs) were selected systematically from the national census frame with probability proportional to size, followed by random selection of households within each EA to ensure national representation.

The analysis used the Individual Recode (IR) dataset of women aged 15–49 years. Sampling weights provided by the DHS Program were applied to account for the survey’s complex design, differential selection probabilities, and nonresponse. After weighting, the final analytical sample comprised 13,951 women. Detailed ZDHS methodology is available from the DHS Program [17].

### Study variables and measurements

#### Study variables

The primary outcome was high-risk fertility behavior (HRFB), defined as maternal age <18 or >34 years, short birth interval (<24 months), or high parity (≥4 births). Women not meeting any criteria were classified as not having HRFB. Explanatory variables included socio-demographic, economic, and reproductive health factors: maternal and partner education, employment, household wealth, residence, media exposure, sex of household head, history of pregnancy termination, contraceptive use, and participation in healthcare decision-making. Media exposure was defined as engagement with radio, television, or newspapers at least once per week; contraceptive use as current use of any modern method; and maternal education as no education, primary, secondary, or higher.

Ethical approval for the ZDHS was obtained by the DHS Program from the relevant national ethics committee and ICF International. No additional approval was required for this secondary analysis of anonymized data.

#### Data Management and Analysis

All analyses applied sampling weights to generate nationally representative estimates. Data cleaning and analysis were conducted in STATA 17. Descriptive statistics summarized socio-demographic and reproductive health characteristics. Given the hierarchical DHS data and high prevalence of HRFB, multilevel mixed-effects Poisson regression with robust standard errors was used to estimate adjusted prevalence ratios (aPR) and 95% confidence intervals, accounting for clustering at the enumeration area level. Variables with p < 0.20 in bivariable analysis or conceptual relevance were included in the multivariable model. Model fit was assessed using deviance, AIC, BIC, intra-class correlation coefficient, and median odds ratio (MOR).

#### Multilevel Analysis of Factors Associated with High-Risk Fertility Behavior

We fitted four hierarchical models to examine determinants of high-risk fertility behavior (HRFB): a null model with no covariates to quantify baseline cluster variation, an individual-level model, a community-level model, and a combined model including both levels. Clustering was assessed using the Likelihood Ratio (LR) test, intra-class correlation coefficient (ICC), and median odds ratio (MOR), indicating the proportion of HRFB variation attributable to differences between clusters. The ICC, reflecting the proportion of total variance arising from between-cluster differences, was computed for logistic and Poisson mixed-effects models as:

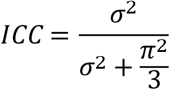 where σ^2^represents the estimated cluster-level variance.

The MOR quantifies unexplained cluster-level heterogeneity by expressing it as an odds ratio, indicating how much the likelihood of HRFB changes when moving from a lower-risk to a higher-risk cluster. The MOR was calculated using:

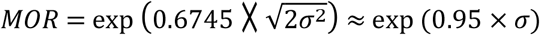

where σ^2^ again denotes the cluster-level variance.

Variables with p < 0.20 in bivariable analyses or considered conceptually important were included in multivariable models. Model fit was assessed using deviance statistics, with the lowest deviance indicating the most optimal model. Final estimates are presented as adjusted prevalence ratios (aPR) with 95% confidence intervals (CI), and predictors with p < 0.05 in the multivariable model were considered statistically significant.

## RESULTS

### Characteristics and Prevalence of High-Risk Fertility Behavior

Table 1 presents the socio-demographic and reproductive characteristics of the 13,951 women included in the analysis, alongside the prevalence of high-risk fertility behavior (HRFB). Most women were aged 15–24 years (41.8%), followed by 25–34 years (30.2%) and 35–49 years (28.0%). Nearly half had completed secondary education (46.5%), while 6.5% had no formal schooling; partners’ education showed a similar pattern. Over half of the women (58.5%) reported no regular media exposure, 38.4% were current users of modern contraceptives, and 12.5% had a history of pregnancy termination. Overall, HRFB was reported by 48.4% of women, increasing sharply with age (22.1% for 15–24 years, 49.3% for 25–34 years, 81.1% for 35– 49 years) and decreasing with higher maternal and partner education. Similar trends were observed for household wealth, media exposure, and women’s participation in healthcare decisions, indicating that HRFB is concentrated among older, less-educated women with limited resources and access to health information.

**Table 1:** Distribution of the study population by socio-demographic and reproductive-related characteristics (n = *13951*)

| Variables | Weighted frequency (%) | High risk fertility behaviour |  |
| --- | --- | --- | --- |
|  |  | Yes (%) | No (%) |
| Individual level variables |  |  |  |
| Age of respondent |  |  |  |
| 15-24 | 5889 (41.81%) | 1381 (22.06) | 4508 (77.94) |
| 25-34 | 4114 (30.24%) | 2119 (49.25) | 1995 (50.75) |
| 35-49 | 3948 (27.95%) | 3259 (81.13) | 689 (18.87) |
| Women education status |  |  |  |
| No education | 959 (6.505) | 717 (75.36) | 242 (24.64) |
| Primary | 5849 (39.28) | 3702 (62.82) | 2147 (37.18) |
| Secondary | 6189 (46.48) | 2121 (33.35) | 4068 (66.65) |
| Higher | 954 (7.734) | 219 (22.18) | 735 (77.82) |
| Husbands' education status |  |  |  |
| No education | 359 (4.679) | 290 (80.96) | 69 (19.04) |
| Primary | 2599 (32.45) | 2004 (77.05) | 595 (22.95) |
| Secondary | 3481 (48.04) | 2084 (58.51) | 1397 (41.49) |
| Higher | 805 (11.96) | 327 (40.23) | 478 (59.77) |
|  |  | Yes (%) | No (%) |
| Don't know | 213 (2.871) | 155 (72.65) | 58 (27.35) |
| Women employment status |  |  |  |
| Not employed | 7440 (53.31) | 3062 (39.24) | 4378 (60.76) |
| Employed | 6511 (46.69) | 3697 (55.42) | 2814 (44.58) |
| Household wealth status |  |  |  |
| Rich | 5531 (46.22) | 1903 (34.3) | 3628 (65.7) |
| Middle | 2839 (19.11) | 1520 (53.56) | 1319 (46.44) |
| Poor | 5581 (34.67) | 3336 (59.71) | 2245 (40.29) |
| Sex of household head |  |  |  |
| male | 9468 (67) | 4685 (47.85) | 4783 (52.25) |
| female | 4483 (33) | 2074 (44.65) | 2409 (55.35) |
| Media exposure |  |  |  |
| no | 8604 (58.54) | 4718 (53.82) | 3886 (46.18) |
| yes | 5347 (41.46) | 2041 (36.87) | 3306 (63.13) |
| Women's healthcare decision making |  |  |  |
| Respondent alone | 2333 (32.52) | 1525 (63.38) | 808 (36.62) |
| Jointly | 3439 (47.28) | 2236 (63.87) | 1203 (36.13) |
| Husband, parent, or<br>someone else | 1568 (20.2) | 1045 (65.8) | 523 (34.2) |
| Current contraceptive use |  |  |  |
| No | 8564 (61.57) | 3330 (36.95) | 5234 (63.05) |
| Yes | 5387 (38.43) | 3429 (62.57) | 1958 (37.43) |
| Ever had a terminated pregnancy |  |  |  |
| No | 12202 (87.51) | 5679 (44.91) | 6523 (55.09) |
| Yes | 1749 (12.49) | 1080 (59.98) | 669 (40.02) |
| **Community level variables** |  |  |  |
| Residence |  |  |  |
| Urban | 6131 (50.79) | 2352 (37.51) | 3779 (62.49) |
| Rural | 7820 (49.21) | 4407 (56.37) | 3413 (43.63) |

### Multilevel Analysis of Factors Associated with High-Risk Fertility Behavior

Table 2 presents the multilevel mixed-effects Poisson regression results for determinants of high-risk fertility behavior (HRFB). Individual-level factors explained most of the variability (full model ICC ≈ 0, MOR = 1). Older maternal age was the strongest predictor, with women aged 25–34 years showing 28% higher prevalence (aPR = 1.282, 95% CI: 1.202–1.367) and those 35–49 years showing 89% higher prevalence (aPR = 1.888, 95% CI: 1.783–1.999) compared with women aged 15–24 years. Secondary (aPR = 0.781,95% CI: 0.738–0.826) and higher education (aPR = 0.530, 95% CI: 0.459–0.612) were protective, as was higher partner education. Employment (aPR = 1.037) and current contraceptive use (aPR = 1.136) increased HRFB, whereas poorer household wealth (aPR = 0.930) and a history of terminated pregnancy (aPR = 0.936) were protective. Rural residence was not significant (aPR = 1.040), indicating that geographic variation is largely explained by individual characteristics.

**Table 2:**
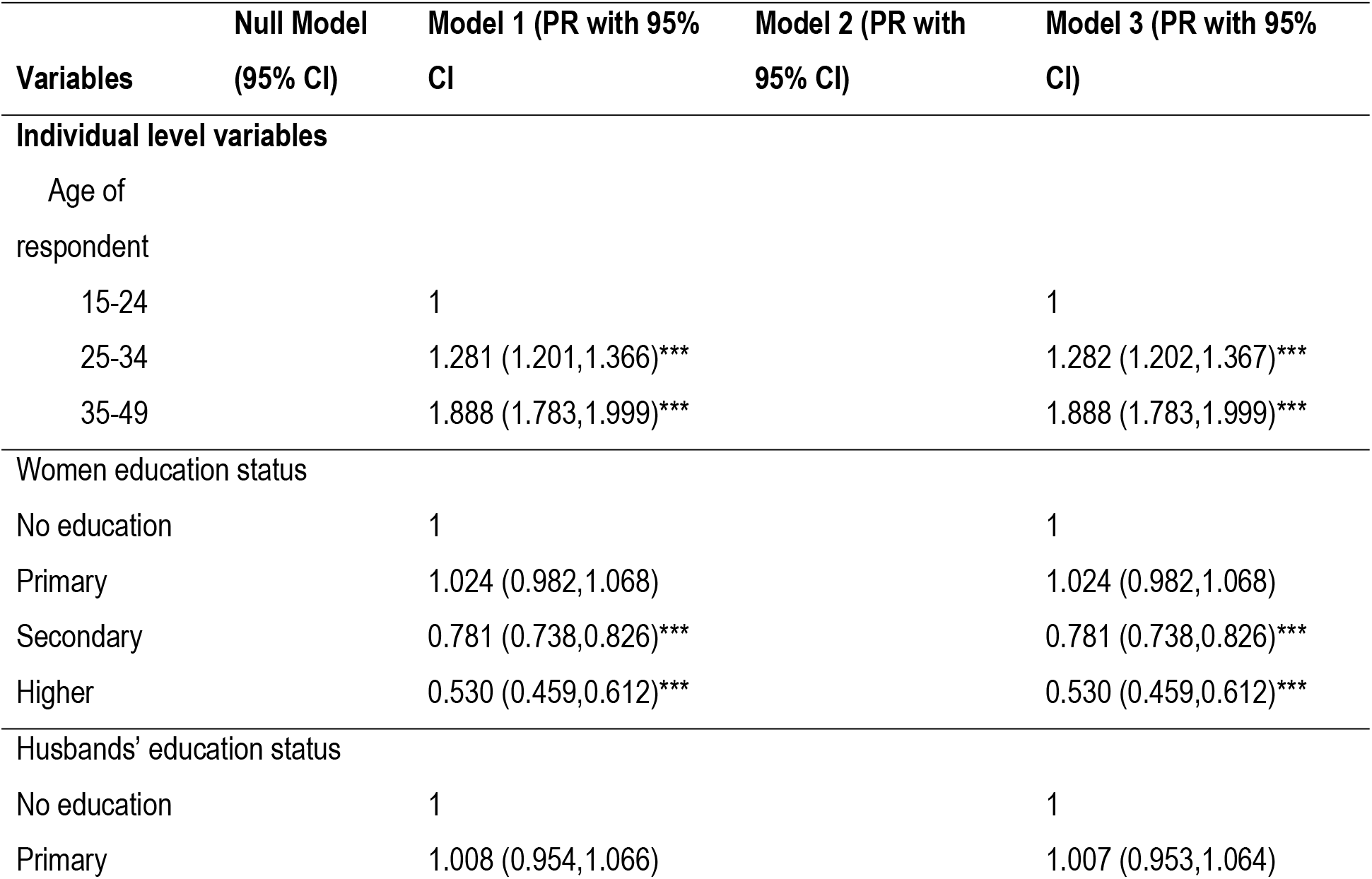
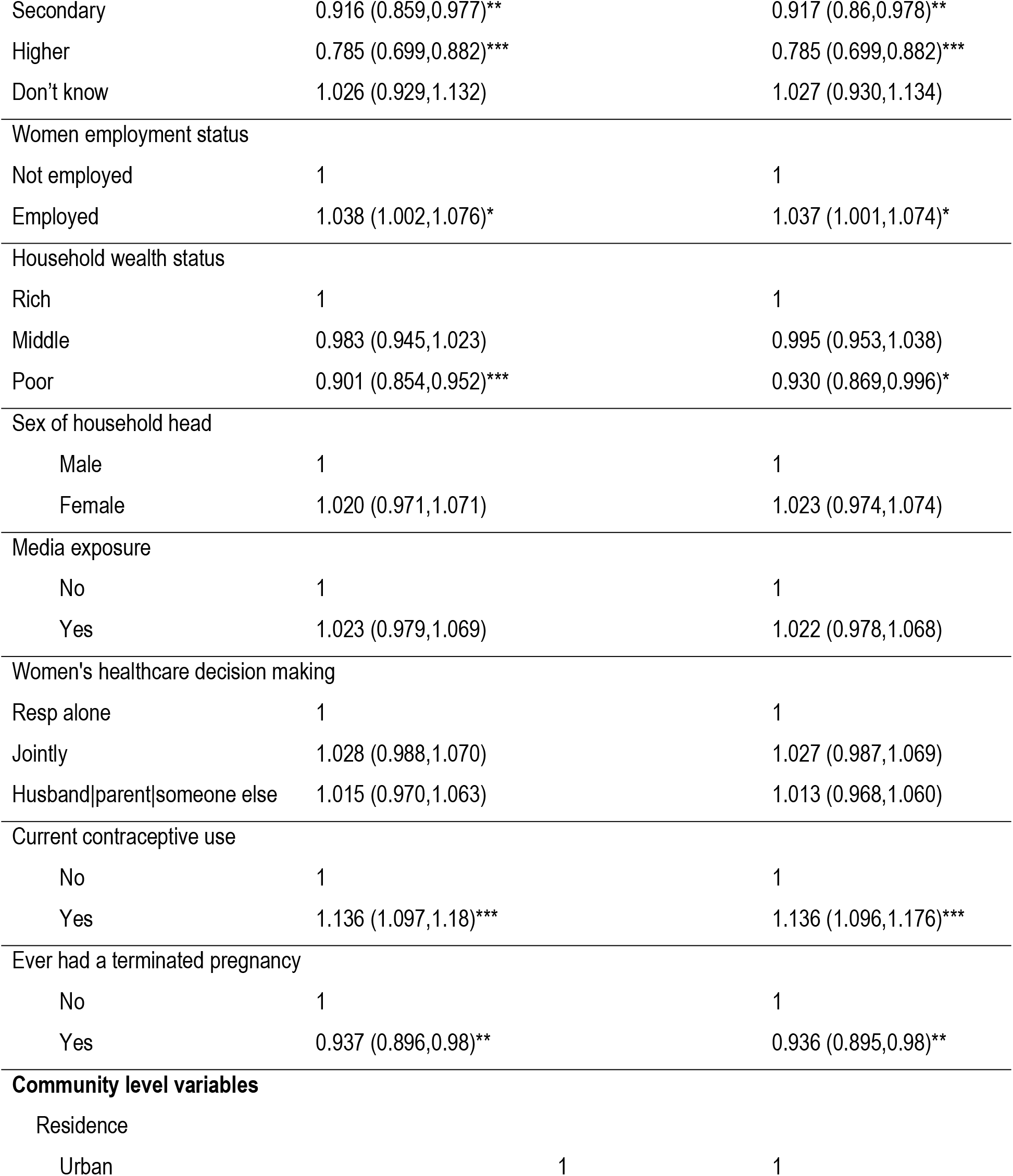
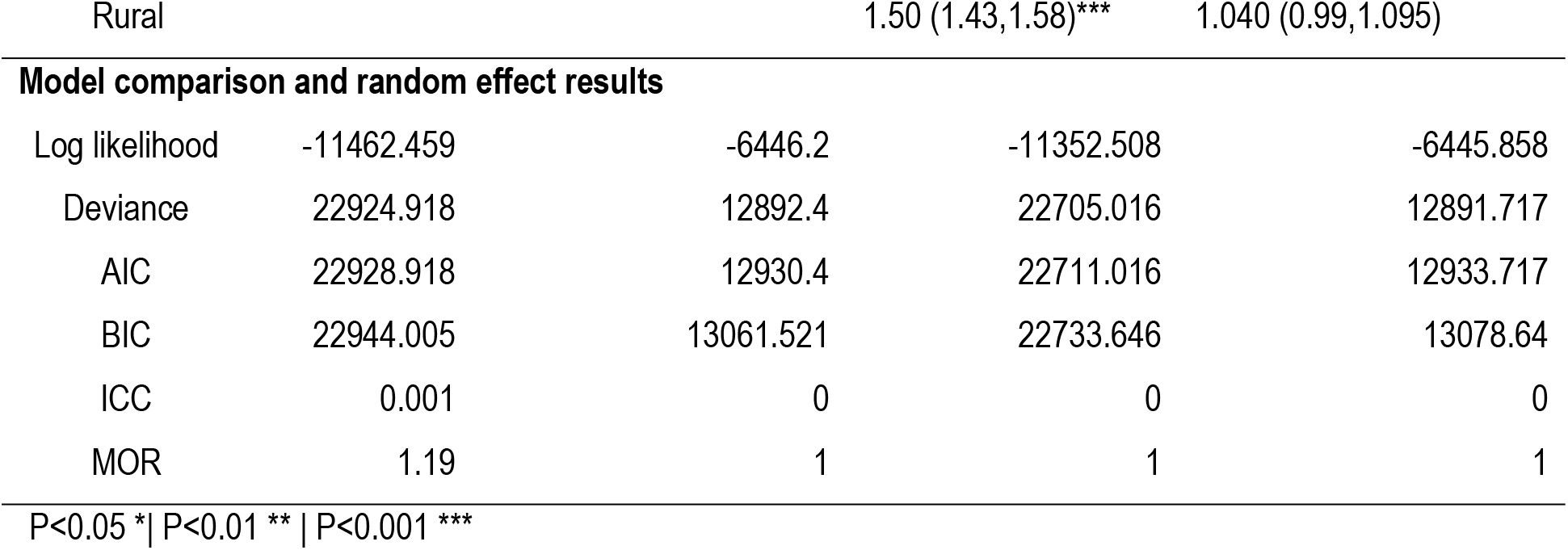
Multilevel Analysis of Factors Associated with High-Risk Fertility Behavior.

| Variables | Null Model<br>(95% CI) | Model 1 (PR with 95%<br>CI) | Model 2 (PR with<br>95% CI) | Model 3 (PR with 95%<br>CI) |
| --- | --- | --- | --- | --- |
| **Individual level variables** |  |  |  |  |
| Age of<br>respondent |  |  |  |  |
| 15-24 |  | 1 |  | 1 |
| 25-34 |  | 1.281 (1.201,1.366)*** |  | 1.282 (1.202,1.367)*** |
| 35-49 |  | 1.888 (1.783,1.999)*** |  | 1.888 (1.783,1.999)*** |
| **Women education status** |  |  |  |  |
| No education |  | 1 |  | 1 |
| Primary |  | 1.024 (0.982,1.068) |  | 1.024 (0.982,1.068) |
| Secondary |  | 0.781 (0.738,0.826)*** |  | 0.781 (0.738,0.826)*** |
| Higher |  | 0.530 (0.459,0.612)*** |  | 0.530 (0.459,0.612)*** |
| **Husbands' education status** |  |  |  |  |
| No education |  | 1 |  | 1 |
| Primary |  | 1.008 (0.954,1.066) |  | 1.007 (0.953,1.064) |

| **Variables** | **Null Model**<br>**(95% CI)** | **Model 1 (PR with 95%**<br>**CI)** | **Model 2 (PR with**<br>**95% CI)** | **Model 3 (PR with 95%**<br>**CI)** |
| --- | --- | --- | --- | --- |
| Secondary |  | 0.916 (0.859,0.977)** |  | 0.917 (0.86,0.978)** |
| Higher |  | 0.785 (0.699,0.882)*** |  | 0.785 (0.699,0.882)*** |
| Don't know |  | 1.026 (0.929,1.132) |  | 1.027 (0.930,1.134) |
| **Women employment status** |  |  |  |  |
| Not employed |  | 1 |  | 1 |
| Employed |  | 1.038 (1.002,1.076)* |  | 1.037 (1.001,1.074)* |
| **Household wealth status** |  |  |  |  |
| Rich |  | 1 |  | 1 |
| Middle |  | 0.983 (0.945,1.023) |  | 0.995 (0.953,1.038) |
| Poor |  | 0.901 (0.854,0.952)*** |  | 0.930 (0.869,0.996)* |
| **Sex of household head** |  |  |  |  |
| Male |  | 1 |  | 1 |
| Female |  | 1.020 (0.971,1.071) |  | 1.023 (0.974,1.074) |
| **Media exposure** |  |  |  |  |
| No |  | 1 |  | 1 |
| Yes |  | 1.023 (0.979,1.069) |  | 1.022 (0.978,1.068) |
| **Women's healthcare decision making** |  |  |  |  |
| Resp alone |  | 1 |  | 1 |
| Jointly |  | 1.028 (0.988,1.070) |  | 1.027 (0.987,1.069) |
| Husband\|parent\|someone else |  | 1.015 (0.970,1.063) |  | 1.013 (0.968,1.060) |
| **Current contraceptive use** |  |  |  |  |
| No |  | 1 |  | 1 |
| Yes |  | 1.136 (1.097,1.18)*** |  | 1.136 (1.096,1.176)*** |
| **Ever had a terminated pregnancy** |  |  |  |  |
| No |  | 1 |  | 1 |
| Yes |  | 0.937 (0.896,0.98)** |  | 0.936 (0.895,0.98)** |
| **Community level variables** |  |  |  |  |
| Residence |  |  |  |  |
| Urban |  |  | 1 | 1 |
| Rural |  |  | 1.50 (1.43,1.58)*** | 1.040 (0.99,1.095) |
| **Model comparison and random effect results** |  |  |  |  |
| Log likelihood | -11462.459 | -6446.2 | -11352.508 | -6445.858 |
| Deviance | 22924.918 | 12892.4 | 22705.016 | 12891.717 |
| AIC | 22928.918 | 12930.4 | 22711.016 | 12933.717 |
| BIC | 22944.005 | 13061.521 | 22733.646 | 13078.64 |
| ICC | 0.001 | 0 | 0 | 0 |
| MOR | 1.19 | 1 | 1 | 1 |

P<0.05 \*| P<0.01 \*\* | P<0.001 \*\*\*

## Discussion

The study provides a comprehensive analysis of the prevalence and determinants of HRFB among women of reproductive age in Zambia, utilizing the most recent 2024 Demographic and Health Survey (DHS) data. The finding that nearly half (48.4%) of Zambian women exhibit at least one HRFB underscores the persistent and substantial public health challenge posed by risky fertility patterns in the country. While this proportion is lower than that reported in Tanzania (71.6%)[5], it still represents a significant population at risk for adverse maternal and neonatal outcomes, reinforcing the urgency of addressing HRFB as a national health priority [4], [18].

Consistent with regional evidence, age emerged as a strong and progressive predictor of HRFB in Zambia. Women aged 25–34 and 35–49 years demonstrated substantially higher prevalence ratios compared to younger women, mirroring trends observed across sub-Saharan Africa where high parity and advanced maternal age are major contributors to HRFB [4], [18], [19]. The accumulation of risk among older women may be attributed to repeated childbearing, shorter birth intervals driven by sociocultural expectations, and delayed or limited access to effective family planning [5], [19], [20]. These findings highlight the need for targeted fertility-limiting interventions and tailored reproductive health services for older women of reproductive age.

Education consistently emerged as one of the most strong protective factors against HRFB. Women with secondary or higher education, as well as those whose husbands had higher educational attainment, showed significantly lower prevalence of HRFB [5], [21], [22]. This aligns with extensive literature demonstrating that education enhances women’s autonomy, expands reproductive choices, and improves communication regarding fertility preferences[18], [22], [23]. Educated women are more likely to delay marriage and childbearing, utilize modern contraceptives, and make informed reproductive decisions, as evidenced in both Zambian and broader East African contexts [19], [23]. These findings underscore the critical importance of investing in female education as a structural determinant of reproductive health.

The relationship between household wealth and HRFB in Zambia revealed complex patterns. Descriptive analyses indicated higher HRFB among poorer women, yet multivariable models showed a modest reduction in risk among the poorest households compared to the wealthiest. This contrasts with findings from Tanzania and Ethiopia, where poverty consistently increased vulnerability to HRFB [18], [19]. The Zambian pattern may reflect behavioral adaptations such as earlier cessation of childbearing among poorer women or more frequent use of extended breastfeeding, which is associated with longer birth intervals [21], [22]. Nevertheless, persistent socioeconomic disparities highlight the need to address barriers to reproductive health services and information among disadvantaged groups

Unexpectedly, contraceptive users in Zambia showed a higher prevalence of HRFB, a finding that aligns with research suggesting that women often initiate contraception after experiencing high-risk pregnancies or reaching high parity [5], [24]. This reactive pattern of contraceptive uptake underscores the need to promote earlier, proactive initiation of family planning, ideally before the accumulation of risk factors. Strengthening preconception counseling and integrating family planning into broader reproductive health services could help shift contraceptive use from a corrective to a preventive strategy [23].

Contrary to evidence from Tanzania and Bangladesh, media exposure and women’s participation in healthcare decision-making did not show significant independent effects on HRFB in adjusted analyses [19], [21], [25]. This suggests that information pathways and decision-making dynamics in Zambia may be more complex, or that media exposure alone is insufficient to drive behavioral change in certain communities. It also highlights the importance of context-specific strategies that go beyond information dissemination to address underlying social and cultural determinants of fertility behavior[25], [26].

The high prevalence of adolescent pregnancy in Zambia (29–48%) further compounds the HRFB burden, with early marriage, limited access to sexual and reproductive health services, and entrenched socio-cultural norms identified as key drivers [3], [21], [27]. These findings reinforce the need for multi-level interventions that address individual, interpersonal, community, and policy-level barriers to delaying childbearing and promoting healthy reproductive trajectories among adolescents.

Rural residence was linked to higher HRFB in the analyses, reflecting well-documented disparities in access to health services, family planning, and education in rural sub-Saharan Africa[3], [5], [28]. The elevated HRFB in rural areas is largely explained is compounded by lower education and socioeconomic status rather than rural location alone. This pattern aligns with recent evidence showing that structural inequities, not geography per se, drive maternal health risks in the region [28].

Women with a history of terminated pregnancy showed a slightly lower prevalence of HRFB. Similar findings suggest that pregnancy loss may prompt women to adopt healthier fertility behaviors, such as increased birth spacing or seeking reproductive health counseling [3], [29]. This highlights the potential for post-abortion care services to serve as critical entry points for promoting healthy fertility planning and reducing future HRFB [29].

This study leveraged recent nationally representative DHS data and robust multilevel analytical methods, providing reliable and generalizable insights into HRFB in Zambia. Key strengths include the large sample and sophisticated modeling approach, which accounted for clustering at the community level. However, the cross-sectional design limits causal inference, and self-reported fertility behaviors may be subject to reporting biases. Furthermore, complex sociocultural influences on fertility may not be fully captured by quantitative measures alone.

The findings highlight several actionable strategies. Programs should promote early family planning uptake, strengthen post-abortion counselling, and provide targeted support for older women continuing childbearing. Investments in girls’ education remain a critical long-term strategy to reduce high-risk fertility patterns. Health communication and reproductive health services should be enhanced to provide clear guidance on birth spacing and timing, empowering women to make informed fertility decisions. Policies prioritizing these individual-level interventions can help reduce maternal and neonatal risks and align Zambia with national and global reproductive health goals.

## Conclusion

HRFB remain common in Zambia, affecting nearly half of women of reproductive age. Older maternal age, lower education, limited access to health information, and cumulative childbearing patterns were the main drivers, while education for women and their partners was strongly protective. Contraceptive use often occurred after risk behaviors were already established. Most variation in high-risk fertility occurred at the individual rather than community level, underscoring the need for interventions that target personal circumstances to improve maternal and newborn health outcomes.

## Data Availability

The data underlying the results presented in this study are available from the Demographic and Health Surveys (DHS) Program. The 2024 Zambia Demographic and Health Survey data are publicly available upon request through the DHS Program website at: https://dhsprogram.com/data/ Access to the data requires user registration and approval by the DHS Program to ensure appropriate use of the data. All analyses were conducted using anonymized, nationally representative survey data provided by the DHS Program.

## Acknowledgements

This work was supported by the Southern Africa Research Capacity Network (SOFAR), funded by the European & Developing Countries Clinical Trials Partnership (EDCTP), grant number 101145636.

## Notes

### Competing Interest Statement

The authors have declared no competing interest.

### Clinical Trial

NA

### Funding Statement

The author(s) received no specific funding for this work.

### Author Declarations

This study utilized de-identified, publicly available data from the 2024 Zambia Demographic and Health Survey (ZDHS). Access to the data was granted through the DHS Program, and, because the dataset is fully anonymized and publicly accessible, no additional ethical approval was required for this secondary analysis.

